# MENDS-on-FHIR: Leveraging the OMOP common data model and FHIR standards for national chronic disease surveillance

**DOI:** 10.1101/2023.08.09.23293900

**Authors:** Shahim Essaid, Jeff Andre, Ian M Brooks, Katherine H Hohman, Madelyne Hull, Sandra L Jackson, Michael G Kahn, Emily M Kraus, Neha Mandadi, Amanda K Martinez, Joyce Y Mui, Bob Zambarano, Andrey Soares

**Affiliations:** Department of Biomedical Informatics, University of Colorado Anschutz Medical Campus, Denver CO; Commonwealth Informatics Inc, Waltham MA; Health Data Compass, University of Colorado Anschutz Medical Campus, Denver CO; National Association of Chronic Disease Directors (NACDD), Decatur GA; National Center for Chronic Disease Prevention and Health Promotion, Centers for Disease Control and Prevention (CDC), Atlanta GA; Kraushold Consulting, Denver CO; Public Health Informatics Institute, Decatur, GA; Department of Medicine, University of Colorado Anschutz Medical Campus, Denver CO

**Keywords:** 1. Health Information Interoperability [L01.470.813], 2. Public Health Surveillance [N06.850.780.675.487], 3. Health Level Seven [N03.540.630.480], 4. Electronic Health Records [N06.850.520.308.940.968.625.250], 5. HL7 Fast Healthcare Interoperability Resources (FHIR)

## Abstract

**Objective:** The Multi-State EHR-Based Network for Disease Surveillance (MENDS) is a population-based chronic disease surveillance distributed data network that uses institution-specific extraction-transformation-load (ETL) routines. MENDS-on-FHIR examined using Health Language Seven’s Fast Healthcare Interoperability Resources (HL7^®^ FHIR^®^) and US Core Implementation Guide (US Core IG) compliant resources derived from the Observational Medical Outcomes Partnership (OMOP) Common Data Model (CDM) to create a standards-based ETL pipeline.

**Materials and Methods:** The input data source was a research data warehouse containing clinical and administrative data in OMOP CDM Version 5.3 format. OMOP-to-FHIR transformations, using a unique JavaScript Object Notation (JSON)-to-JSON transformation language called Whistle, created FHIR R4 V4.0.1/US Core IG V4.0.0 conformant resources that were stored in a local FHIR server. A REST-based Bulk FHIR $export request extracted FHIR resources to populate a local MENDS database.

**Results:** Eleven OMOP tables were used to create 10 FHIR/US Core compliant resource types. A total of 1.13 trillion resources were extracted and inserted into the MENDS repository. A very low rate of non-compliant resources was observed.

**Discussion:** OMOP-to-FHIR transformation results passed validation with less than a 1% non-compliance rate. These standards-compliant FHIR resources provided standardized data elements required by the MENDS surveillance use case. The Bulk FHIR application programming interface (API) enabled population-level data exchange using interoperable FHIR resources. The OMOP-to-FHIR transformation pipeline creates a FHIR interface for accessing OMOP data.

**Conclusion:** MENDS-on-FHIR successfully replaced custom ETL with standards-based interoperable FHIR resources using Bulk FHIR. The OMOP-to-FHIR transformations provide an alternative mechanism for sharing OMOP data.

**LAY ABSTRACT:** Many chronic conditions, such as hypertension, obesity, and diabetes are becoming more prevalent, especially in high-risk individuals, such as minorities and low-income patients. Public health surveillance networks measure the presence of specific conditions repeatedly over time, seeking to detect changes in the amount of a disease conditions so that public health officials can implement new early-prevention programs or evaluate the impact of an existing prevention program. Data stored in electronic health records (EHRs) could be used to measure the presence of health conditions, but significant technical barriers make current methods for data extraction laborious and costly. HL7 BULK FHIR is a new data standard that is required to be available in all commercial EHR systems in the United States. We examined the use of BULK FHIR to provide EHR data to an existing public health surveillance network called MENDS. We found that HL7 BULK FHIR can provide the necessary data elements for MENDS in a standardized format. Using HL7 BULK FHIR could significantly reduce barriers to data for public health surveillance needs, enabling public health officials to expand the diversity of locations and patient populations being monitored.

## 1 INTRODUCTION

The COVID-19 pandemic highlighted the urgent need for rapid access to clinical data from diverse settings to assess emerging risk factors and treatment outcomes [1–6]. Public health networks that collect, harmonize, and report on acute diseases have traditionally relied on manual health surveys and local data collection methods. Quickly expanding these networks to a national scale that reaches a wide range of populations and settings has significant technical and sustainability challenges [6,7]. Similarly, chronic disease surveillance registries need timely access to linked clinical, administrative, and social determinants of health data across diverse healthcare settings [8]. Chronic disease surveillance must address additional challenges, given the need for diagnostic, therapeutic, and observational longitudinal data over many years. Although electronic health record (EHR) systems capture detailed longitudinal data in health-seeking populations, these data are difficult to extract and harmonize to common data structures and terminologies [9–13].

The Multi-State EHR-Based Network for Disease Surveillance (MENDS) pilot project focuses on harmonizing clinical data from EHRs to support chronic disease monitoring at scale and across disparate clinical settings [14]. Focusing on data elements and measures related to hypertension, smoking, statin use, diabetes, and obesity, MENDS aims to inform local and national health departments regarding the chronic disease burden and outcomes at the population level. The MENDS data infrastructure is built using Electronic Medical Record Support for Public Health (ESP), an open-source software suite [15–18]. A detailed description of the MENDS governance, technical structure, and data elements has been published previously [19,20].

Health Level Seven’s Fast Healthcare Interoperability Resources (HL7^®^ FHIR^®^) is an extensive international health data exchange standard based on units of data exchange, called Resources, that must conform with explicit standards for structure (data formats), content (allowed terms), and operations (queries, updates, data exchanges) [21]. Commercial implementation of FHIR-based interfaces and applications has increased dramatically [22]. In addition to responding to traditional marketplace forces, in the United States, EHR software vendors must comply with the 21st Century Cures Act, which contains regulatory mandates with implementation deadlines, certification criteria, and penalties for non-conformance requiring implementation of FHIR Version 4.0 and US Core Implementation Guideline (US Core IG) Version 4.0.0 by December 31, 2022 [23,24].

Two additional elements of the FHIR specification are FHIR Profiles [25] and Implementation Guides (IGs) [26]. FHIR Profiles define specifications that narrow or expand the scope of a FHIR base resource definition. For example, a Profile can specify additional data fields, alter fields from optional to mandatory, define relationships between data elements, or declare alternative or expanded terminologies or value sets in a FHIR resource. An IG is a collection of Profiles that defines a specific use case for a resource. The US Core IG is widely deployed in the United States because it is closely aligned to the data domains and terminologies defined in the US Core Data for Interoperability (USCDI) Version 1. USCDI V1 defines the set of mandatory elements for data exchange required by legislation to certify commercial EHR systems.

FHIR-based data exchange occurs using two basic models: real-time single-patient and batch-oriented bulk data queries. Both standards use the same data and coding formats (FHIR Resources, Profiles, IGs) but Bulk FHIR returns data for all patients in a cohort in a single asynchronous batch-oriented operation [27]. Bulk FHIR is designed to enable population-focused use cases, such as public health surveillance, clinical quality assessment, and health services research [28]. The Bulk FHIR standard, which is in early development, is less widely deployed than single-patient FHIR. Mandatory conformance certification for Bulk FHIR export standards was required by December 31, 2022, while Bulk FHIR import standards have been delayed [29,30]. However, interest in Bulk FHIR is growing and a few Bulk FHIR public health applications have been developed [31]. For example, VACtrac is a public health use case that uses Bulk FHIR to exchange vaccine data between health institutions and a state immunization registry [32]. In addition, Jones et al. compared the Bulk FHIR export capabilities of several commercial and open-source FHIR servers [33].

Currently, MENDS data are imported using several custom extraction-transformation-load (ETL) processes. Detailed specifications describe the format and content for each MENDS data element. MENDS data contributors write custom ETL routines, involving significant technical burden [11]. An alternative ETL approach for MENDS that uses FHIR Resources combined with the US Core IG and HL7 Bulk FHIR export could significantly reduce the technical effort by enabling access to standardized data that are independent of underlying database structures and supported by commercial EHR vendors. This pilot project was devised to test this hypothesis.

In seeking data partners for a Bulk FHIR pilot, MENDS discovered that production-ready EHR Bulk FHIR interfaces are not yet widely available or, if provided by the EHR vendor, are not supported in operational heath system IT environments. An alternative approach to pilot testing Bulk FHIR as a standards-based ETL method for MENDS was devised. Patient-level clinical data stored in an Observational Medical Outcomes Partnership common data model (OMOP CDM) within a research data warehouse (RDW) setting was transformed into standardized FHIR Resources and loaded into a separate dedicated commercial FHIR server. The clinical data contained in the OMOP CDM was sufficient to create the set of FHIR resources needed by the MENDS database. This pilot FHIR server effectively emulated the anticipated state of an EHR-based FHIR server without requiring health system resources.

This report presents one technical approach for enhancing interoperable data exchange in chronic disease surveillance efforts. This implementation also can inform others how to leverage Bulk FHIR-based data exchange, especially institutions with access to relevant clinical data stored in the OMOP CDM format. The report describes a pipeline that transforms OMOP data into US Core IG compliant FHIR resources, uploads the FHIR resources into a FHIR server, provides FHIR output in response to a Bulk FHIR request to the FHIR server, and uploads the resulting FHIR resources into the MENDS database. Technical results include the size of the data exchange this pipeline generated for the MENDS use case and estimates of the elapsed time to transform OMOP to FHIR, import FHIR Bundles into a FHIR server, export Bulk FHIR from a FHIR server, and upload the Bulk FHIR files into the MENDS database as an example of real-world experience with this approach.

## 2 METHODS

The originating data source is a research data warehouse store in an OMOP CDM Version 5.3 relational database hosted by Health Data Compass (HDC). HDC is a research data custodian for clinical, administrative, genomic, and external data sources sponsored by the University of Colorado Anschutz Medical Campus, UCHealth, Children’s Hospital Colorado, and University Physicians Inc (http://healthdatacompass.org). HDC’s technical infrastructure is hosted in the Google Cloud Platform (GCP). All HDC databases are instances of GCP’s BigQuery enterprise data management environment [34]. Figure 1 is a Level 1 logical data flow diagram illustrating the data processing pipeline.

**Figure 1.**
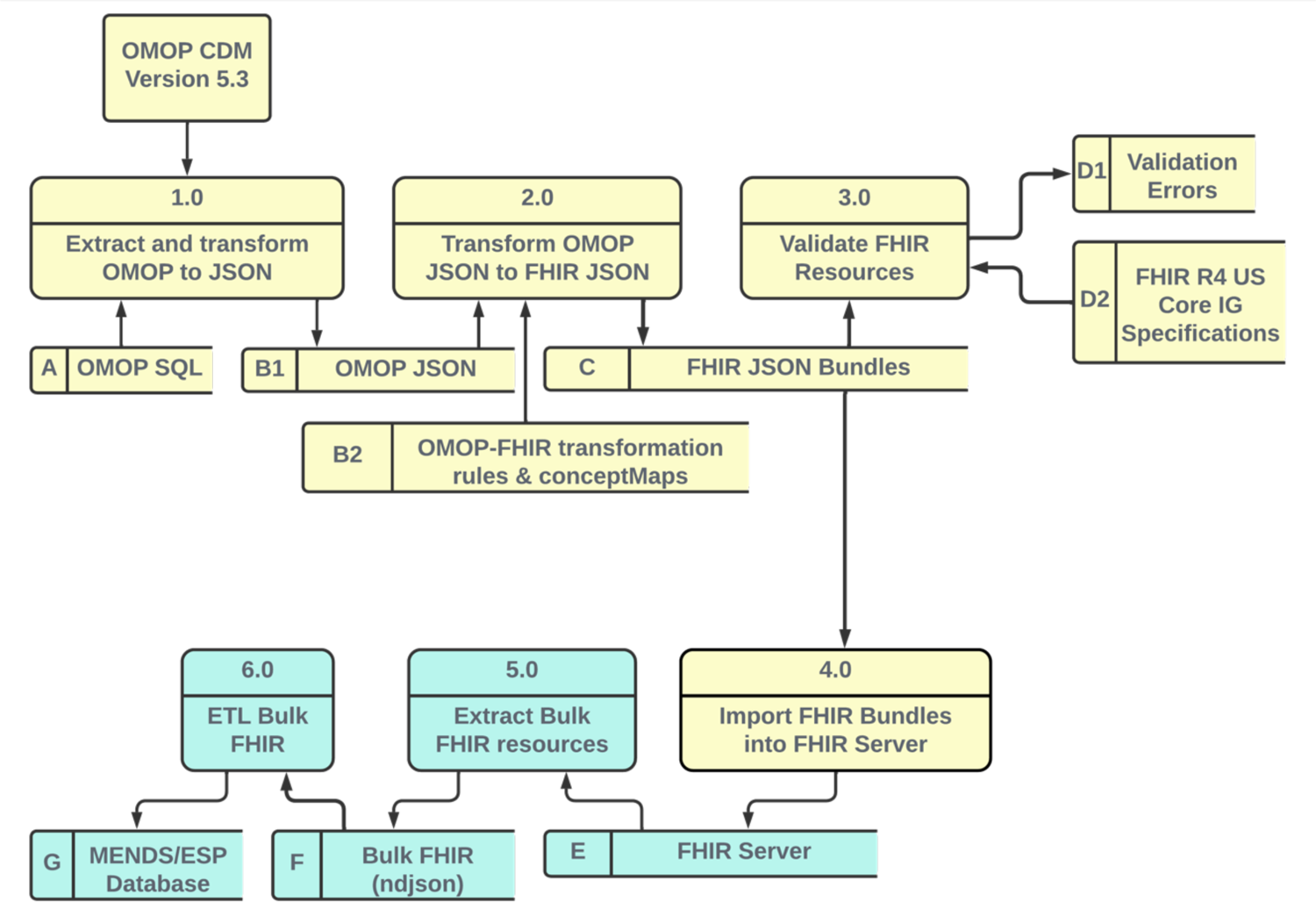
Level 1 logical data flow diagram. Yellow components perform the OMOP-to-FHIR transformation and FHIR server upload. Green components perform the Bulk FHIR export and FHIR-based ETL into the MENDS database. [Figure 1_MENDS_Level_1_Logical_Dataflow.png]

The yellow data flow begins with clinical data extracted from UCHealth’s Epic^®^ EHR stored in a GCP BigQuery relational data mart in OMOP CDM V5.3 format. The MENDS data mart cohort are all patients seen at UCHealth who met the following criteria:

- Must have at least one “clinical” visit (inpatient, outpatient, or emergency department) on or after January 1, 2017
- Must have age >=2 years
- Must have a 5 digit zip code and state of residence (MENDS mandatory variables)

A full OMOP data extract, called the MENDS data mart, was created for all patients in the MENDS cohort.

The MENDS data mart is transformed into a set of US Core IG V4.0.0 compliant FHIR resources and loaded into a FHIR server. This is the OMOP-to-FHIR transformation phase. The green data flow extracts Bulk FHIR resources and inserts the resulting extracted FHIR resources into the MENDS/ESP database. This is the pilot FHIR-to-MENDs ETL phase. A version of the OMOP-to-FHIR transformation and FHIR server import (Steps 2.0–4.0) with synthetic data in OMOP JavaScript Object Notation (JSON) format is freely available on GitHub (https://github.com/CU-DBMI/mends-on-fhir).

### 2.1 Dataflow 1.0: OMOP-to-OMOP JSON

ANSI-standard SQL statements query OMOP CDM V5.3 tables to generate output rows that create FHIR resources. All data elements required for a FHIR resource must be included in the SQL output. For example, the OMOP Death table is LEFT JOINed with the OMOP Person table so that a death date, when available, is included in the query results. Each SQL statement creates an “OMOP JSON” object (Figure 1, B1) consisting of a single key and an array. The JSON key represents the original OMOP table. Each element in the JSON array is an OMOP database row. Figure 2 illustrates the OMOP JSON output for the OMOP Condition_Occurrence table.

**Figure 2.**
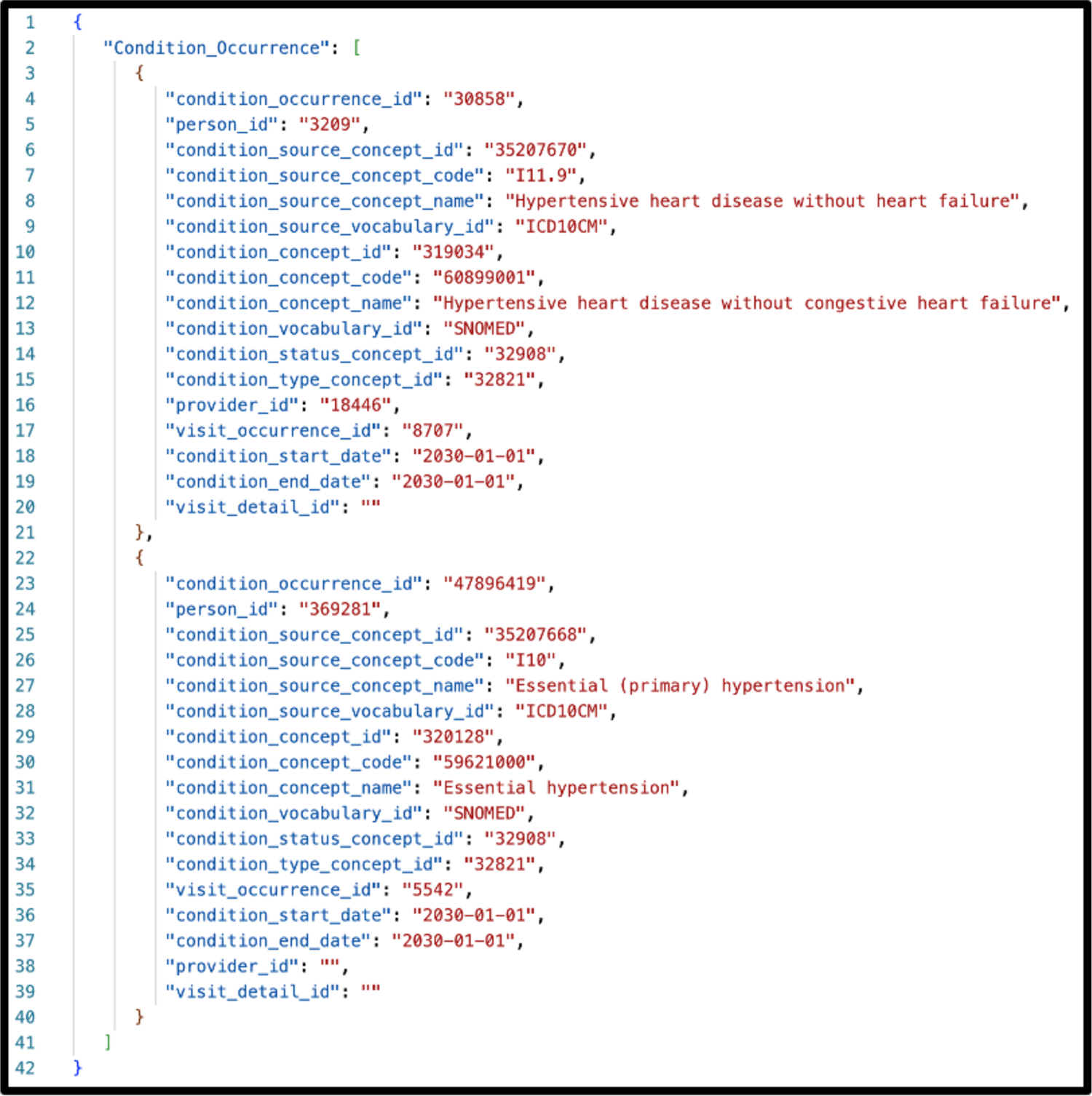
Structure of “OMOP JSON” extracted from OMOP CDM V5.3 queries using synthetic data based on the OMOP Condition_Occurrence table. The person_id, provider_id, and condition_ start/end dates fields do not refer to actual values. [Figure2_OMOP_JSON.png]

### 2.2 Dataflow 2.0: OMOP JSON to FHIR R4 Bundle JSON (OMOP-to-FHIR)

OMOP JSON is transformed into FHIR R4 Bundle JSON format using an open-source JSON-to-JSON transformation engine that implements a functional language called Whistle [35]. Whistle transformation files create FHIR output that is conformant to FHIR R4 V4.0.1 and US Core IG 4.0.0 specifications. Figure 3 illustrates a portion of the Whistle specification for transforming an OMOP PERSON record into a FHIR PATIENT resource. Elements on the left side of a colon are either internal variables or FHIR JSON keys. Elements on the right side of a colon are OMOP fields, i.e., Whistle functions that modify OMOP fields, or constants. The transformation functions are applied to each row in the OMOP JSON array, creating one or more FHIR resources. A terminal function wraps the array of FHIR resources into a single FHIR Bundle resource.

**Figure 3.**
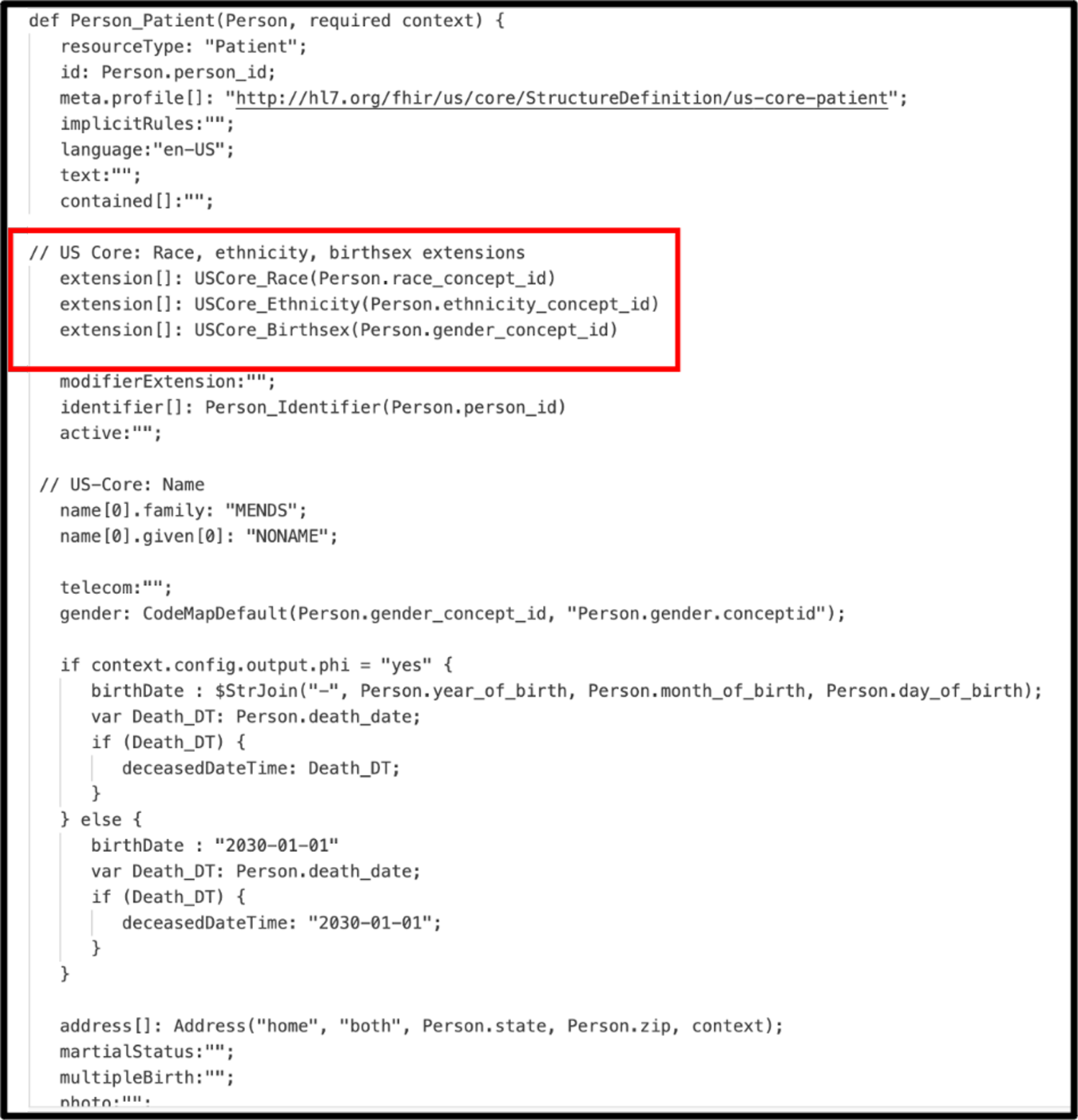
Whistle transformation specification for creating FHIR R4 Person resource from OMOP CDM V5.3 Patient record. Functions such as USCore_Birthsex() use a unique feature of the Whistle transformation language that calls FHIR concept maps to convert OMOP-specific concept_ids into US Core IG-compliant CodeableConcepts. [Figure3_Whistle_Transformation.png]

One unique feature of the Whistle mapping language is a built-in function focused on code harmonization using local FHIR ConceptMap resources or remote FHIR terminology services. For example, the Whistle function USCore_Birthsex() in Figure 3 (red box) uses the local FHIR ConceptMap shown in Figure 4 to translate OMOP-specific concept_ids into US Core IG conformant values. Owing to Internet access restrictions, the implementation only uses local ConceptMaps for all OMOP-to-FHIR terminology mappings.

**Figure 4.**
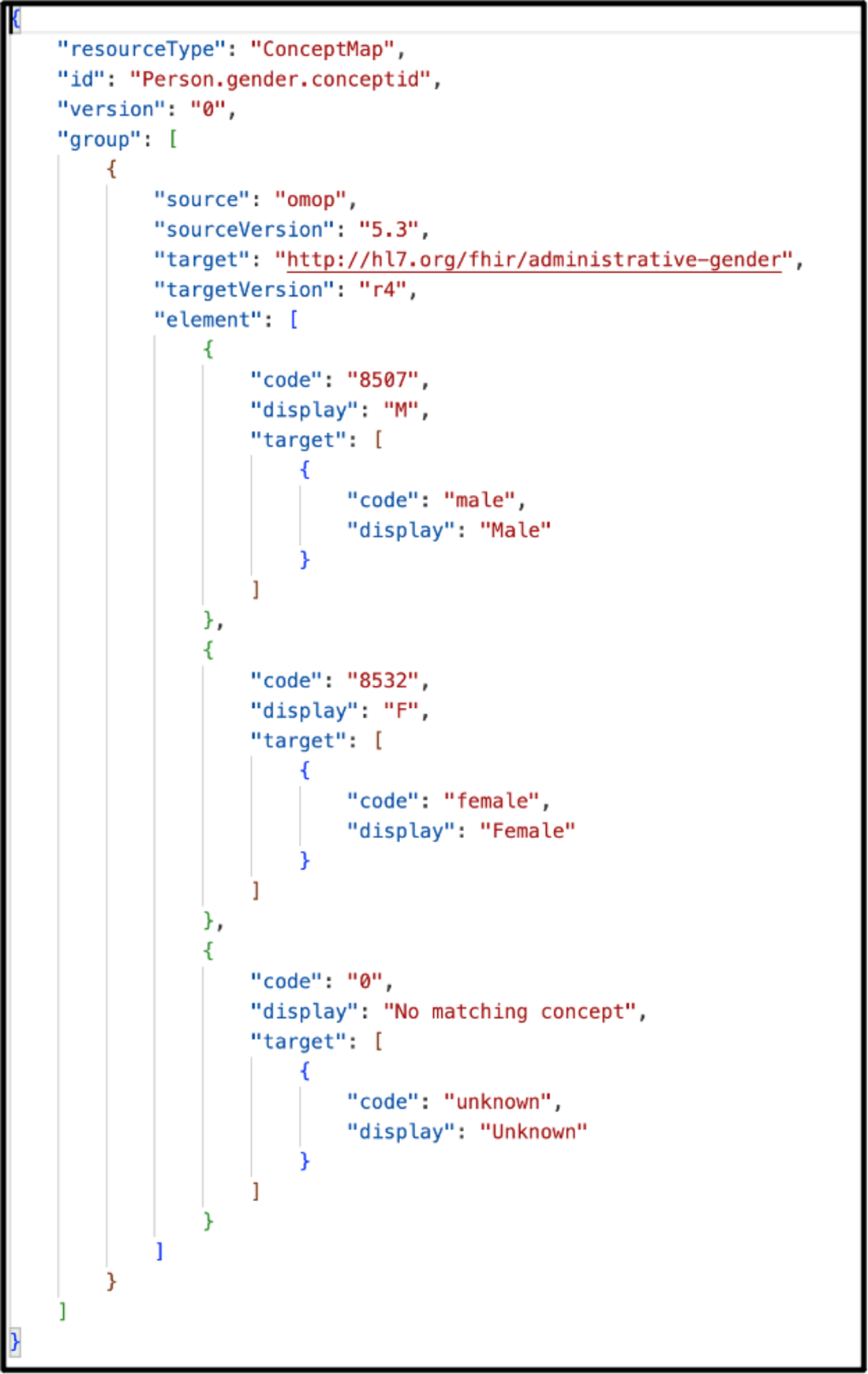
FHIR ConceptMap maps OMOP-specific concept_ids for patient sex into FHIR US Core compliant values. [Figure4_FHIR_ConceptMap.png]

MENDS-on-FHIR resources also includes nonstandard original EHR source values and codes. Figure 5 illustrates the inclusion of both the U.S. Core mandated RxNorm code mapped from the drug_concept_id (red box) and the nonstandard National Drug Code (NDC) code (green box) from the EHR in a FHIR Medication resource. The same approach enabled FHIR Condition resources to contain both nonstandard ICD9CM/ICD10CM source codes along with US Core-required Systematized Nomenclature of Medicine (SNOMED) codes.

**Figure 5.**
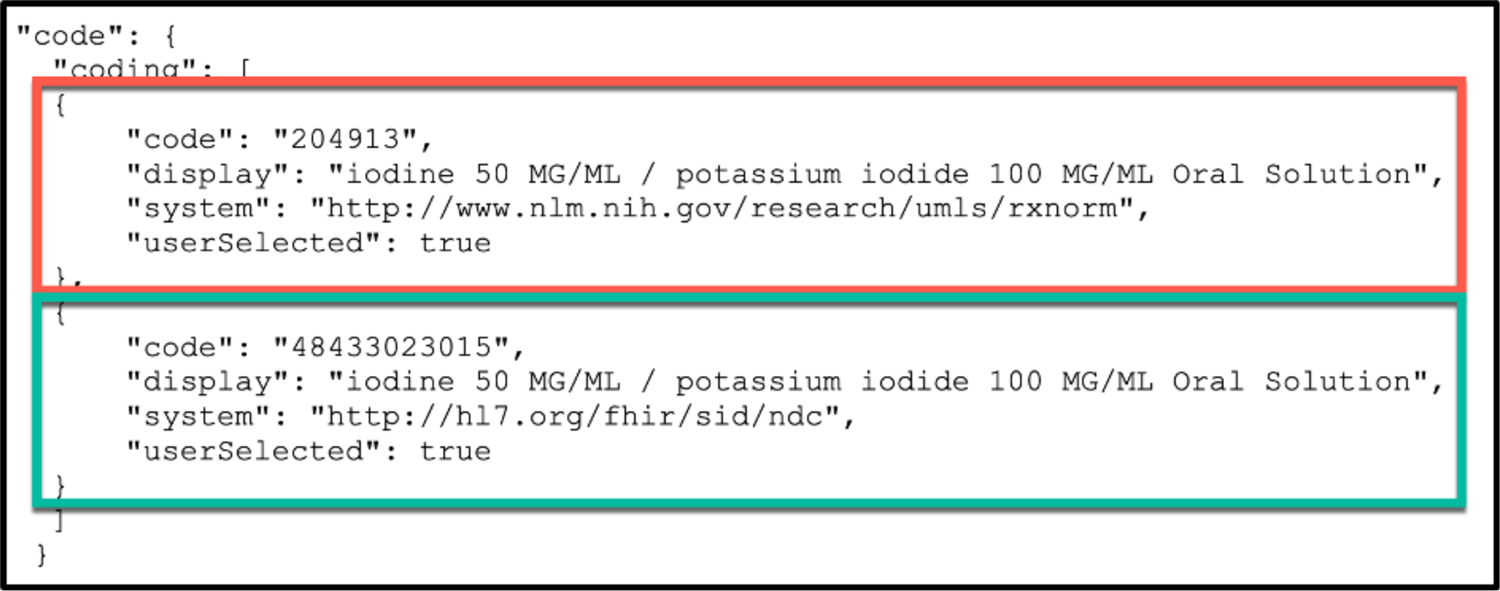
Multiple JSON Codings in a FHIR code element enable inclusion of both local source and FHIR-required values. In this example, both FHIR-required RxNorm (red box) and local NDC source codes (green box) are included in two Coding objects in the Medication.code JSON object. [Figure5_MultipleCodings.png]

### 2.3 Dataflow 3.0: FHIR R4 and US Core IG Validation

FHIR validation tools examine the structure and content of FHIR resources for conformance to FHIR Profiles and IGs using the open-source HL7 FHIR Validator [36]. The validator was configured to use FHIR R4 V4.0.1 and US-Core IG V4.0.0.

### 2.4 Dataflow 4.0: Bulk FHIR Import

Each FHIR Bundle JSON file in Figure 1 C is uploaded to a FHIR server configured with base FHIR R4 and the US Core IG v4.0.0 using a FHIR $import call (Figure 1 Process 4.0). The FHIR server does not perform a referential integrity check at the time of data import; instead, server-generated Resource IDs are created. Import errors are logged for inspection after all resources are loaded. A full data refresh is performed with each FHIR $import.

### 2.5 Dataflow 5.0: Bulk FHIR Extract

The ESP server requests FHIR resources by executing a FHIR $export call to the FHIR server, which launches an asynchronous export process. All instances of a FHIR resource type (Patient, Condition, MedicationRequest, Medication, Immunization, Observation) are exported in one large NDJSON file.

### 2.6 Dataflow 6.0: ESP FHIR Import

After completing the Bulk FHIR extract, ESP triggers a process that imports Bulk FHIR extracts into the MENDS/ESP database.

## 3 RESULTS

Table 1 shows the alignment among the clinical and demographic data domains required by the MENDS database, the FHIR resources that contain these data elements, and the OMOP table(s) used to construct the FHIR resources. The 10 FHIR resources needed by MENDS required data extracted from 10 OMOP tables plus the OMOP CONCEPT table for codes and text labels. OMOP Observation rows were transformed into one of three different FHIR Observation Profiles defined by the US Core IG: Observation (Smoking), Observation (Non-Smoking), and Observation (Laboratory). Three separate transformations were used to create Profile-conformant variants of the Observation Resource. Although the OMOP CDM can map to additional FHIR resources, only those required to meet MENDS chronic disease surveillance use cases were created.

**Table 1.**
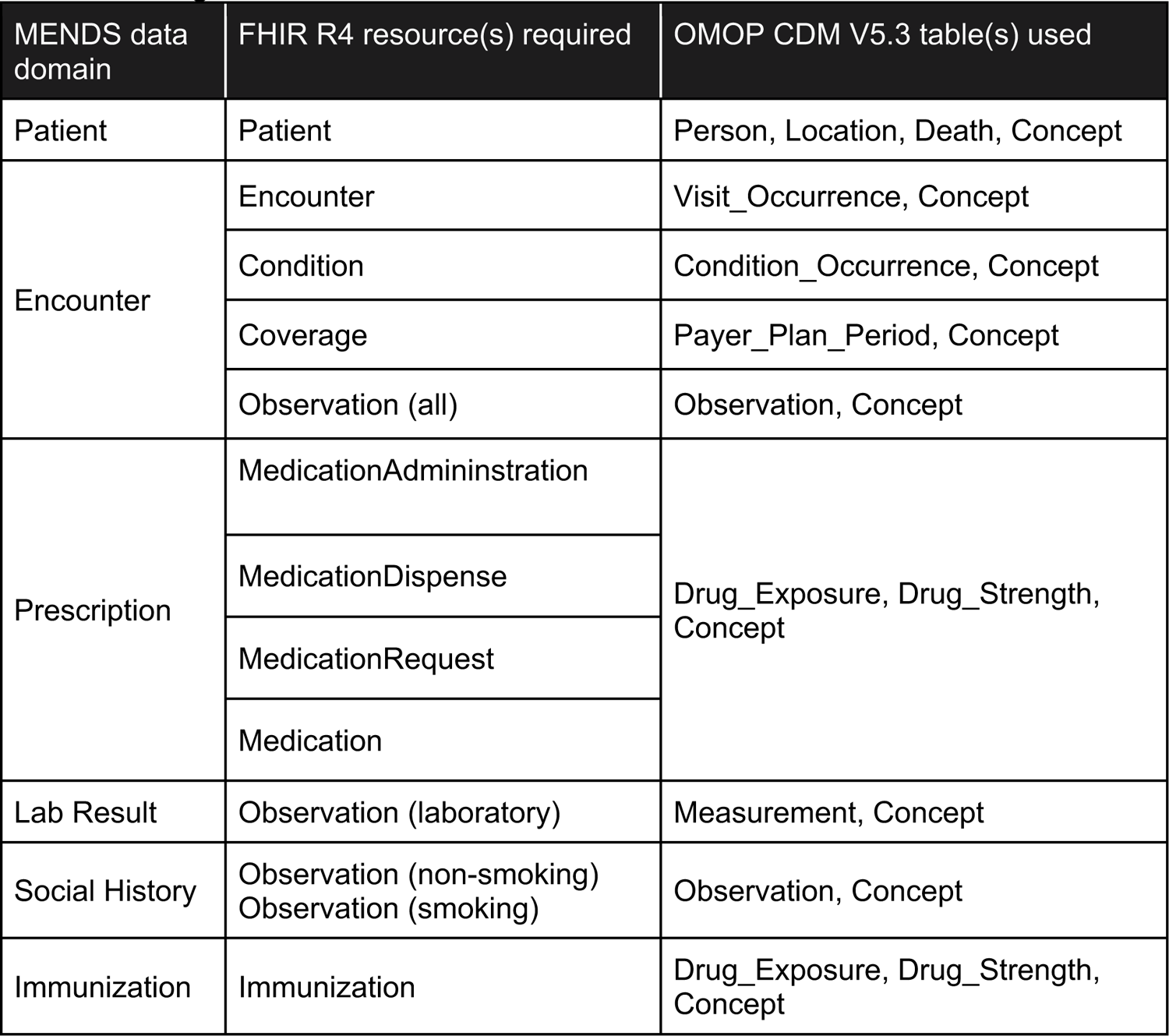
Alignment of MENDS, FHIR R4, and OMOP data models.

| MENDS data domain | FHIR R4 resource(s) required | OMOP CDM V5.3 table(s) used |
| --- | --- | --- |
| Patient | Patient | Person, Location, Death, Concept |
| Encounter | Encounter | Visit_Occurrence, Concept |
|  | Condition | Condition_Occurrence, Concept |
|  | Coverage | Payer_Plan_Period, Concept |
|  | Observation (all) | Observation, Concept |
| Prescription | MedicationAdministration | Drug_Exposure, Drug_Strength, Concept |
|  | MedicationDispense |  |
|  | MedicationRequest |  |
|  | Medication |  |
| Lab Result | Observation (laboratory) | Measurement, Concept |
| Social History | Observation (non-smoking)<br>Observation (smoking) | Observation, Concept |
| Immunization | Immunization | Drug_Exposure, Drug_Strength, Concept |

Table 2 provides basic statistics about the MENDS cohort and the FHIR resources generated using the pilot OMOP CDM. From a total OMOP CDM patient population of 4.38 million, 3.24 million met the very broad MENDS cohort inclusion criteria. For many FHIR resources, there was a one-to-one correspondence between the number of OMOP rows and FHIR resources. OMOP observations and measurements were both represented as FHIR Observation resources, whereas OMOP drug exposure records were distributed across five different medication-related FHIR resources.

**Table 2.** OMOP row counts versus FHIR Resources. OMOP and FHIR counts are for the MENDS cohort only (M=million).

| OMOP |  | FHIR |  |
| --- | --- | --- | --- |
| Table Name | OMOP Rows | FHIR Resource Type | FHIR Resources Generated |
| Person | 3.24M | Patient | 3.24M |
| Visit_Occurrence | 141M | Encounter | 141M |
| Condition_Occurrence | 189M | Condition | 189M |
| Payer_Plan_Period | 162M | Coverage | 162M |
| Observation (smoking)<br>Observation (non-smoking) | 138M | Observation | 411M |
| Measurement (labs) | 399M |  |  |
| Drug_Exposure | 221M | MedicationAdministration | 102M |
|  |  | MedicationDispense | 8.4M |
|  |  | MedicationRequest | 109M |
|  |  | Medication | 55K |
|  |  | Immunization | 640K |

Table 3 provides approximate timing results derived from multiple end-to-end executions for a complete data refresh across the entire 3.24 million patient MENDS cohort.

**Table 3.** Sample processing times (execution times are approximate based on multiple execution runs).

|  | Processing step | Execution time |
| --- | --- | --- |
| OMOP to FHIR Server<br>(Figure 1, yellow objects) | OMOP to OMOP JSON export (Figure 1 Process 1.0) | ~30 minutes |
|  | OMOP JSON to FHIR server import (Figure 1 Processes 2.0 and 4.0) * | ~39 hours |
| FHIR Server to MENDS DB<br>(Figure 1, green objects) | Bulk FHIR export (Figure 1 Process 5.0) | ~30 minutes |
|  | FHIR export to MENDS/ESP import (Process 6.0) | ~80 hours |
| * Process 3.0 (validation) is not performed during production runs. It is only performed as part of Whistle transformation testing. |  |  |

## 4 DISCUSSION

The MENDS-on-FHIR pilot illustrates how Bulk FHIR access to US Core IG conformant FHIR resources can support a large-scale population-level chronic disease surveillance database. MENDS-on-FHIR creates FHIR resources using the OMOP CDM (Figure 1; yellow components). The OMOP CDM contains sufficient detailed patient-level clinical data to meet the MENDS data requirements. The same ESP Bulk FHIR import interface (Figure 1; green components) could be used in a health system’s operational EHR-based FHIR servers when regulatory mandates result in broader deployment. Of note, exporting more than 1.1 billion FHIR resources consumed only approximately 30 minutes (Table 3, Process 5.0). This is the only process in the pipeline that executes on the operational FHIR server in a healthcare setting. This finding suggests that concerns regarding the impact of Bulk FHIR extracts on operational FHIR server performance may not be as significant as expected.

A secondary benefit of this pilot is enabling data access to OMOP data via a Bulk FHIR interface (Figure 1; yellow components). The OHDSI community, which supports the OMOP CDM, currently contains “over 3,260 collaborators in 80 countries across 21 time zones in 6 continents” (https://ohdsi.org/who-we-are/collaborators/; accessed 15-November-2023). A FHIR data exchange capability opens this expansive network to even more data sharing possibilities beyond OMOP-specific queries.

Propelled by regulatory mandates and certification requirements, FHIR data exchange capabilities are present in nearly all U.S. commercial EHR systems. Data aggregators do not have the same regulatory pressures and therefore have been slower to incorporate FHIR capabilities. Although they have implemented FHIR importing functions to consume new data sources, aggregators generally have not developed FHIR exporting functions to share interoperable data with others. External data reporting requirements that use FHIR-based query tools, such as electronic clinical quality measurement reporting, may provide the impetus for data aggregators to add Bulk FHIR export features [37,38]. MENDS-on-FHIR demonstrates one technical approach for adding FHIR capabilities to a clinical data warehouse.

The Whistle language enabled implementation of OMOP-to-FHIR transformations as a batch conversion using a functional JSON-to-JSON programming model. Other batch-oriented OMOP-to-FHIR conversion programs include the original Google Data Harmonization proof-of-concept project [35], UNC CAMP FHIR [39], and FHIR-Ontop-OMOP [40]. VACtrac [32] performed batch conversion of HL7 Immunization messages rather than OMOP as its data source. An alternative approach, using dynamic real-time conversion during query execution, has been implemented by OMOP-on-FHIR [41,42]. Boussadi *et al*. created a similar dynamic FHIR conversion program using the i2b2 CDM as the underlying data source [43]; Kasthurirathne *et al*. used OpenMRS [44]. A batch conversion process does not incorporate new data additions or updates that occur between batch runs. However, a batch transformation, once completed, does not incur transformation overhead during query execution or data extraction. The processing overhead in dynamic translation may go unnoticed when extracting data for a single patient (e.g., mobile apps). However, when executing a query that extracts and transforms multiple FHIR resources in a large cohort as illustrated in Table 2, dynamic transformation is simply not tenable.

A second distinction is the choice of data transformation languages. Whistle is a template-based functional JSON-to-JSON conversion language that allows transformation functions to be combined into higher level functions. Whistle implements functions that operate on FHIR ConceptMaps to support terminology mappings.

MENDS-on-FHIR created functions for US Core IG compliant Code mappings, manipulating Coding arrays, mapping Code_Systems, converting measurement units, and reformatting Date and DateTime fields into FHIR standard formats. Whistle transformations are stored in configuration-like text files rather than being embedded in program code.

### 4.1 Limitations and Challenges

MENDS-on-FHIR limited the scope of OMOP-to-FHIR transformations to OMOP data required to meet the MENDS data requirements and conform to the US Core IG. The only conformant extension was including local source codes when allowed by the FHIR specification. Including source values aided debugging when original provenance was needed.

#### 4.1.1 Data model challenges

OMOP, FHIR, and ESP define data elements and allowed values differently. Thus, translating from OMOP into FHIR and then into ESP entails a field-by-field analysis to identify how a field required by ESP could be represented in FHIR and found in OMOP (backwards translation). With any data translation, mapping fidelity is a concern [45], although mapping errors may have smaller-than-anticipated impact on analytic results [46–48]. For example, OMOP associates insurance coverage over an interval of time that is not tied to clinical events. ESP links a primary payer to each clinical encounter. FHIR defines a Coverage Resource that directly maps to the OMOP interval-based representation. The ESP FHIR importer converts FHIR Coverage Resource intervals into an encounter-based representation using a configurable hierarchy to select a primary payer when there are overlapping coverage periods.

#### 4.1.2 Mapping challenges

Incomplete transformations occurred when field values could not be directly aligned between OMOP and FHIR. Inferred values were used when semantically justified. Otherwise, field values were left blank, even if this decision caused validation errors.

#### 4.1.2.1 Medications

Some mandatory FHIR data elements do not have an equivalent data value from OMOP and were inferred. For instance, the FHIR MedicationRequest.status was set to “stopped” if the OMOP Drug_Exposure.stop_reason was present and set to “unknown” otherwise. For MedicationAdminstration.status and MedicationDispense.status, if an end date existed, the status was considered “completed”, or otherwise “in-progress”.

The MedicationRequest.requester is a mandatory data element, but this information is not always present in the OMOP Drug_Exposure table. When absent, the Drug_Exposure.provider_id was used. If both were absent, the requester field was left blank. When the requester field is blank, the HL7 Validator correctly identifies the resource as being US Core IG non-conformant. Although non-conformant, these resources are still stored in the FHIR Server and are available for Bulk FHIR extracts.

FHIR medication-related resources contain a doseQuantity field to record the amount of a medication per dose. OMOP has defined methods for calculating drug dose from the medication ingredient (https://ohdsi.github.io/CommonDataModel/drug_dose.html). However, due to the complexity of calculating drug doses for multi-ingredient medications, doseQuantity in a FHIR resource is included only when the OMOP Drug_Exposure.quantity field is available. Future work is needed to properly calculate drug dosages in multi-ingredient medications.

##### 4.1.2.2 Smoking

OMOP smoking information represents a class of clinical data where multiple OMOP rows represent answers to a survey instrument. In the HDC OMOP CDM, up to 10 rows were entered from the smoking survey. The FHIR transformer requires all information about a resource to be available in a single row. For the FHIR smoking observation resource, a SQL query was created that concatenated all responses to the smoking questionnaire in an encounter into a single “source value” string. For example, nine separate smoking responses were concatenated into a single “value” that was used to determine the correct SNOMED code in the FHIR smoking observation resource.

*(Chew:No) – (CigarettePacksPerDay:<20) – (Cigarettes:No) – (Cigars:No) – (Nicotine dependence, cigarettes, uncomplicated) – (Pipes:No) – (Snuff:No) – (TobaccoUse:Yes) – (TobaccoUseInYears:+)*

The combination of smoking responses generated thousands of unique combinations. While SNOMED-CT contains several dozen smoking-related codes, the US Core IG only allows six valid codes. MENDS-on-FHIR maps the concatenated strings to one of the six valid FHIR codes and also keeps the concatenated “value” in the CodableConcept.text field to retain the raw survey selections made by a patient.

#### 4.1.3 Execution challenges

Execution challenges are issues that arise only during the creation of the FHIR resources in a production environment.

##### 4.1.3.1 Memory

The Whistle Transformation Engine reads and transforms the entire OMOP JSON file into memory before writing the final transformed FHIR structure. Thus, the execution environment requires sufficient memory to hold the OMOP JSON file plus the output FHIR Bundle resource. The Python program that creates OMOP JSON accepts a parameter, called CHUNKSIZE, that partitions the OMOP query results into separate OMOP JSON files containing exactly the CHUNKSIZE number of OMOP rows. This variable is adjusted according to the memory size of each processing node instantiated in the parallel processing pipeline.

##### 4.1.3.2 Validation

Several validation issues were identified that could not be rectified using existing OMOP data, FHIR ConceptMaps, or Whistle transformations. Table 4 lists these unresolved validation errors. Collectively, the error rate was less than 1%.

**Table 4.** FHIR/US Core IG validation errors that could not be resolved.

| Validation issue | Explanation and examples |
| --- | --- |
| Error: Element cardinality | “MedicationRequest.requester” is a required element in US-Core. In rare cases, the OMOP PROVIDER_ID field was blank. By design, a “fake” entry was not created to meet the cardinality requirement. |
| Error: Code not from code system | A small number of valid SNOMED-CT codes were flagged as not valid by the terminology server. On manual verification, these were found to be US-only SNOMED-CT codes. Examples include condition and smoking related codes. Similarly, a small number of valid RxNorm codes were not recognized as RxNorm codes by the terminology service. Manual verification showed them to be influenza or remapped codes. |
| Error: Violations of FHIR invariant rules) | On very rare occasions, OMOP data contained datetime values where the start date was later than the end value in the Medication.validityPeriod field. These resources were kept “as is” because this is an accurate representation of the original data and has no impact on the current use case. |
| Warning: Code not from value set | US-Core binds the “Medication.code” element to a “Medication Clinical Drug” value set derived from RxNorm. Validation indicated RxNorm codes that did not belong to this value set. Manual examination revealed valid RxNorm codes for the package form, but the value set did not include package-related codes. |
| Warning: No coding from the value set | Validation indicated certain records did not contain any RxNorm codes when one was required. These Drug_Exposure records contained HCPCS (J1200) and OMOP RxNorm extension codes (OMOP1088103), CVX, CPT but no RxNorm code. |
| Warning: Label not matching from terminology server | The validator verifies code label text and warns if there is not an exact match with the label string in the terminology service. A few of the OMOP concept labels had variations from the original labels, such as additional spaces around hyphens. Also, OMOP truncates all labels at 256 characters. |
| Info: Unknown extensions | A FHIR Bundle extension was added to hold the OMOP-specific metadata found in the OMOP CDM_SOURCE table. This extension was correctly flagged as an unknown extension. |
| Info: Unknown code systems | Terminologies not known to the terminology server but that are still assigned URLs by HL7 are present in OMOP source values and source codes. The data elements holding these codes could not be verified because the code system is not available at the terminology server. Examples are HCPCS, OMOP RxNorm extension, and CPT. |

One additional validation challenge involved the inability to detect very low frequency errors using small test sets during validation testing (10,000 resources per resource type). Low frequency errors were detected only at runtime when the full data set was processed. Thus, resources that passed validation testing still had rare runtime errors.

### 4.2 Future Work

MENDS-on-FHIR limits the scope of FHIR resources to include only those required by the MENDS project and the FHIR/US Core IG. The OMOP CDM contains clinical and administrative data across a broad range of data domains, such as clinical procedures, devices, and notes, that could create more FHIR resources.

Even within the included domains, the OMOP CDM has many data elements that MENDS-on-FHIR does not use. For example, the OMOP Drug_Exposure table includes data on patient-informed medications, which could be used to create FHIR MedicationStatement resources. Another opportunity for future expansion is immunization records. FHIR specifies immunization data coded in the CVX CodeSystem, but OMOP allows immunizations to also be coded using the RxNorm CodeSystem. The current Immunization Whistle transformation template only includes CVX records. Mapping RxNORM immunization records into CVX is possible with the existing OMOP Concept_Relationship table but was not done with the current immunization transformation template.

## 5 CONCLUSION

The MENDS-on-FHIR pipeline makes two related but distinct contributions:

- Replacing an institution-specific custom CSV-based data import with US Core IG compliant FHIR resources and Bulk FHIR data access demonstrates the viability of using existing FHIR standards to support a national chronic disease public health surveillance use case. Observed timings suggest Bulk FHIR extracts may not consume significant FHIR server resources.
- Transforming the OMOP research data warehouse into US Core IG compliant FHIR resources expands standards-based data access methods to research data.

Both contributions add to the growing landscape of interoperable data exchange using FHIR-based standards. Using Bulk FHIR as a standards-based data source for population-level surveillance could greatly expand the reach of public health use cases as certified EHR systems meet the 21st Century Cures Act requirements. Linking a FHIR-based interface to an EHR could also improve data timeliness.

The OHDSI community has established practices that enable data sharing across OMOP sites using OMOP-specific tools. Enabling access to OMOP data via FHIR resources expands data access options for OMOP data use in broader settings. For example, tools for executing and visualizing population-specific clinical quality measures using FHIR resources is an area of active development that could leverage OMOP data via a Bulk FHIR interface. Given the high level of innovation and commercial activities using FHIR-based data access, providing FHIR access to OMOP data increases an institution’s return-on-investment in implementing and maintaining this international RDW data asset.

## Data Availability

Software source code with synthetic data available on GitHub

https://github.com/CU-DBMI/mends-on-fhir

https://github.com/CU-DBMI/mends-on-fhir-example-data

## ACKNOWLEDGMENTS

The authors acknowledge the contribution of the MENDS partner sites and project team that participated in the creation of the MENDS data network (https://chronicdisease.org/page/MENDSinfo/). The authors also acknowledge the open-source contribution by the Google Healthcare Data Harmonization proof of concept project [35], which created the Whistle transformation engine and example templates. HL7® and FHIR® are the registered trademarks of Health Level Seven International and use of these trademarks does not constitute an endorsement by HL7.

## 6 STATEMENT OF AUTHORS CONTRIBUTIONS

SE, JA, MH, MGK, NM, BZ, and AS developed the software and GitHub site. SE, MGK, AS, KHH, SLJ, and AKM created the initial draft manuscript. IMB, MH, EMK, JYM, and BZ provided additional input to subsequent manuscript versions. All co-authors approved the final manuscript prior to submission.

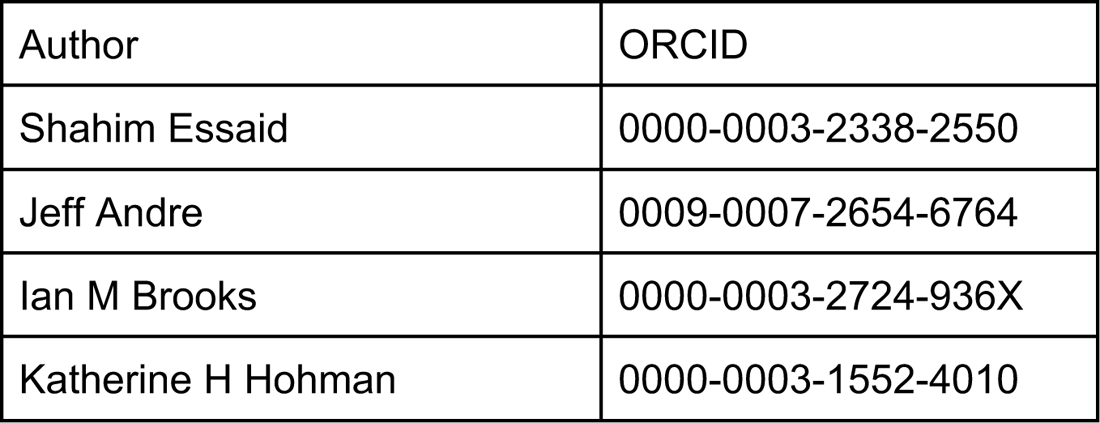
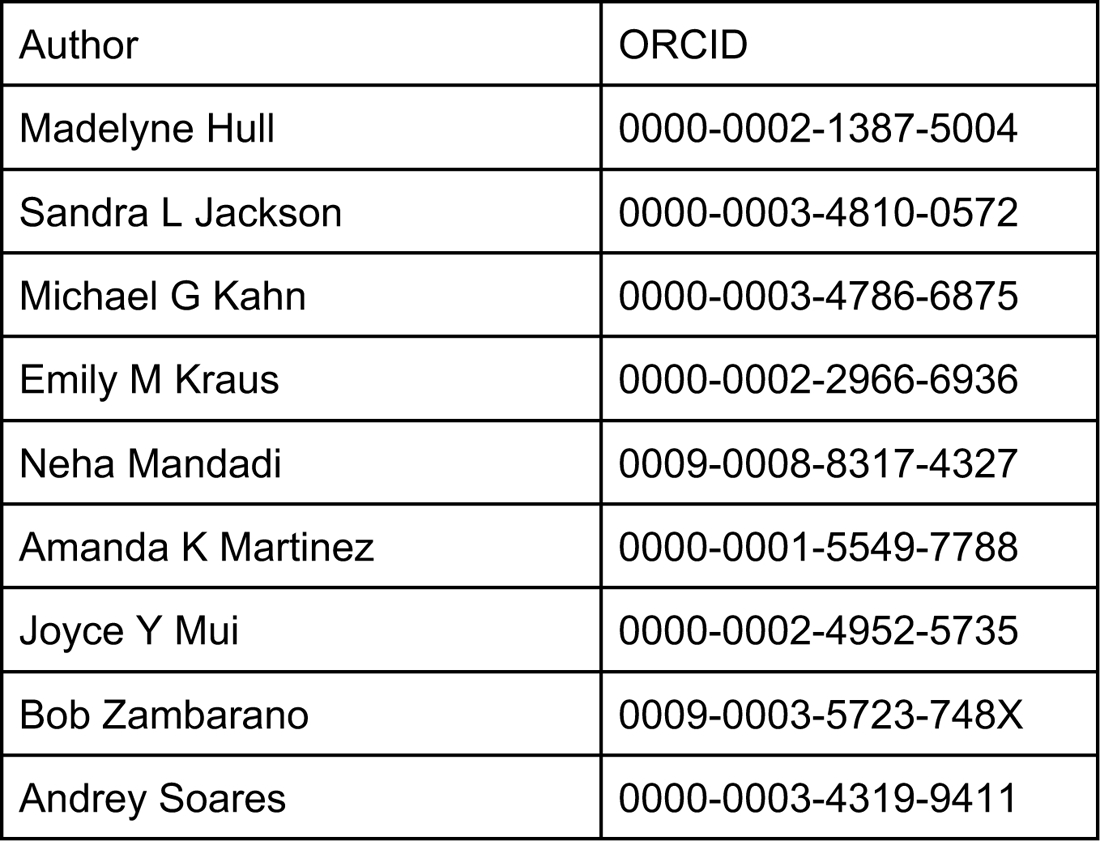

## 7 HUMAN PARTICIPANT COMPLIANCE STATEMENT

CDC provided a written determination that MENDS operates within the public health authority pursuant to the Health Insurance Portability and Accountability Act. As a public health surveillance project, MENDS does not require institutional review board approval.

